# Thymus Composition Predicts Pneumonitis Risk in Lung Cancer Therapy

**DOI:** 10.1101/2025.10.27.25338565

**Authors:** Tafadzwa L. Chaunzwa, Gokul Krishnan, Kwadwo Amoako-Boadu, Abraham J. Book, David G. Miller, Biniam Garomsa, Yuzhong J. Meng, Elaine Yang, Alexis Chidi, Jennifer Ma, Jamie E. Chaft, Narek Shaverdian, Daniel R. Gomez

**Author notes:** Corresponding author: Tafadzwa L. Chaunzwa, MD, DABR, Assistant Member, Department of Radiation Oncology, Memorial Sloan Kettering Cancer Center; Advanced Computing and Oncology Laboratory, Mortimer B. Zuckerman Research Center, 417 East 68th Street, New York, NY 10065.

## Abstract

**Background:** Durvalumab consolidation after concurrent chemoradiotherapy (cCRT) is the standard-of-care for unresectable stage III non–small cell lung cancer (NSCLC) without actionable driver mutations. However, pneumonitis remains a dose-limiting toxicity that often precludes or interrupts immunotherapy. Conventional predictors based on lung dosimetry alone exhibit limited individual-level discrimination. We investigated whether the thymus, long considered vestigial in adults, influences post-treatment immune recovery and susceptibility to inflammatory toxicity.

**Methods:** We analyzed patients with locally advanced NSCLC treated with cCRT in the RTOG 0617 trial (n = 490) and with standard-of-care cCRT followed by consolidation durvalumab at Memorial Sloan Kettering Cancer Center (MSKCC, n = 230). Percent thymic tissue (pTT), a novel imaging parameter that quantifies the proportion of residual functional thymic tissue on pre-treatment CT scans, was derived using an autosegmentation and Gaussian mixture modeling framework. Logistic regression with restricted cubic splines assessed associations between pTT, mean radiation dose to the thymic region (MDTR), and volume of the lungs receiving ≥20 Gy (lung V20) with high-grade (≥ 3) pneumonitis.

**Results:** Across both cohorts, pTT was inversely associated with severe pneumonitis independent of lung V20. Grade 3+ pneumonitis incidence in low-vs high-pTT groups was 7.3% vs 2.9% (p = 0.038) in the RTOG 0617 cohort and 13.9% vs. 5.2% (p = 0.042) in the MSKCC cohort. The combination of low pTT and high lung V20 was associated with the highest rates of severe pneumonitis with 11.5% in RTOG 0617 and 20.8% in MSKCC (compared with 3.3% and 1.9% for high pTT/low lung V20), identifying a subgroup at particularly high risk. MDTR showed a weaker, non-linear relationship with pneumonitis risk, increasing at moderate doses and declining at the highest radiation exposures.

**Conclusions:** This study established pTT, a quantitative marker of residual functional thymus in adults, as a novel, independent predictor of severe pneumonitis following cCRT with or without consolidation immunotherapy. Incorporating pTT into multimodal risk-stratification frameworks could improve patient selection, personalize radiation planning, and enhance safe delivery of curative-intent cCRT and immunotherapy in locally advanced NSCLC.

## INTRODUCTION

The PACIFIC trial transformed treatment for unresectable stage III non–small cell lung cancer (NSCLC) by demonstrating markedly improved survival with durvalumab consolidation after concurrent chemoradiotherapy (cCRT)^1–3^. This regimen, building upon the previous cCRT paradigm established by RTOG 9410 and RTOG 0617 ^4,5^, substantially prolonged median progression-free survival (PFS) and overall survival (OS). Subsequent trials, including PACIFIC-2 and CheckMate 73L, evaluated cCRT with both concurrent and adjuvant immune checkpoint blockade^6,7^, but failed to demonstrate improved outcomes. Thus, the PACIFIC regimen remains the standard of care in unresectable locally advanced NSCLC without actionable driver mutations^1–3,8^. However, this regimen is also associated with a number of treatment-related toxicities, most notably pneumonitis, which can preclude the initiation or continuation of durvalumab. Up to half of patients who receive cCRT are unable to receive or complete consolidation durvalumab due to early disease progression, immune-related/radiation pneumonitis, or other cCRT-associated toxicities^7^.

Pneumonitis is a dose-limiting toxicity in thoracic radiotherapy, associated with significant morbidity and mortality^9,10^. Despite established dose constraints, conventional dosimetric parameters often fail to predict individual risk, with marked variability in toxicities among patients receiving similar lung doses^11,12^. The addition of consolidation immunotherapy further complicates risk assessment. Radiation pneumonitis follows a cascade initiated by pneumocyte injury and dysregulated cytokine signaling, leading to fibrotic remodeling^13^. In contrast, immunotherapy can induce pneumonitis through distinct mechanisms, including T-cell hyperactivation, autoantibody formation, and cytokine imbalance^14,15^. The PACIFIC trial reported a modest increase in pneumonitis rates with durvalumab (<1% increase in severe cases compared to placebo), suggesting predominantly additive rather than synergistic toxicity^2,16^. However, differentiating between radiation, immune-mediated, and radiation recall pneumonitis^17^ is challenging, and misclassification is common, potentially leading to unnecessary treatment discontinuation and loss of therapeutic benefit^18^.

These challenges underscore the need for reliable predictors of pneumonitis during cCRT for locally advanced NSCLC. Although predictive models for pneumonitis have been developed by integrating radiomics, dosiomics, and other biomarkers^10,19–28^, their clinical utility remains limited by variability, lack of standardization, and limited interpretability^29,30^. An emerging area of research has identified the thymus, long regarded as vestigial in adults and routinely excluded from radiation planning, as a potential immunologic regulator in thoracic malignancies treated with radiation and immunotherapy^31^. Radiation to the thymus has been linked to lymphocyte depletion ^32^, while preserved thymic function and regenerative capacity have correlated with improved T-cell reconstitution and immunotherapy response ^33–36^.

Recent studies have generated computed tomography (CT)-based metrics of thymic health and have correlated these metrics with age-related thymic involution and oncologic outcomes^31,37–40^. Building on a mathematical framework introduced by Okamura *et al.*^37,38^, our group developed pTT (percent thymic tissue), a novel imaging biomarker that enables standardized, quantitative assessment of thymic composition across diverse patient populations^31^. We hypothesized that both residual functional thymus, as quantified by pTT, and ionizing radiation dose to the thymic region are associated with pneumonitis risk after cCRT. Specifically, we postulated that lower pTT would correlate with a higher incidence of severe pneumonitis due to immune dysregulation post-cCRT, while higher thymic radiation dose would correlate with an increased incidence of severe pneumonitis due to direct thymic injury or indirectly through concomitant increased lung dose.

To test these hypotheses, we analyzed two distinct cohorts of patients with locally advanced NSCLC treated with cCRT. The RTOG 0617 cohort, treated with cCRT alone, represents a setting in which pneumonitis may be attributable solely to radiation. The Memorial Sloan Kettering Cancer Center (MSKCC) cohort, treated per the PACIFIC regimen with cCRT followed by durvalumab, reflects modern practice with the added potential of immune-related contributions to toxicities. Across both cohorts, pTT emerged as a robust predictor of pneumonitis independent of lung dose, identifying a novel imaging biomarker of pneumonitis risk in patients receiving cCRT with or without durvalumab for NSCLC. In contrast, thymic radiation dose was only associated with pneumonitis risk when analyzed in combination with pTT, suggesting that baseline thymic composition, rather than thymic dose alone, is a key determinant of pneumonitis risk.

## METHODS

### Study population

We retrospectively analyzed data for patients treated for locally advanced NSCLC enrolled in the RTOG 0617 trial^5,41^ and at MSKCC. Participants in the RTOG 0617 cohort received cCRT and were randomized in a 2 x 2 factorial design to radiation prescription doses of 60 Gy versus 74 Gy and cetuximab versus no cetuximab. Patients in the MSKCC cohort were treated according to the PACIFIC regimen, the current standard of care, consisting of cCRT followed by consolidation durvalumab.

### Definition of the thymic region and auto-segmentation

The thymic region was delineated as previously described^31^ using an automated, human-in-the-loop workflow. Briefly, manual contours were generated on radiation planning CTs by a board-certified thoracic radiation oncologist and a senior radiation oncology resident. These annotations were then used to build a deep learning-based autosegmentation model. The thymic region was anatomically defined as the potential space containing residual thymic tissue, bounded superiorly by the level at which the left brachiocephalic vein crosses midline, anteriorly by the sternum, laterally by the mediastinal pleura, and inferiorly by the bottom of the arch of the right pulmonary artery. Automatically generated contours underwent final visual quality assurance. A novel thymic composition and immuno-structural parameter, pTT, was derived from these segmentations and studied for its association with pneumonitis

### Derivation of percent Thymic Tissue (pTT)

A histogram of Hounsfield unit (HU) values within the contoured thymic region on CT was computed for each patient, as previously described^31^, using a mathematical framework first introduced by Okamura et al. and refined to capture components of the thymic space. HU values were binned into 50 equal-width intervals over the entire dynamic range. Only finite HU values were retained, and no other thresholding was used. The histogram was normalized to derive the empirical probability distribution. We then fit a Gaussian mixture model (GMM) to the distribution, and the parameters of the GMM model were estimated using the expectation-maximization (EM) algorithm. We denote *p̂*(*x*) as the empirical distribution obtained from the histogram of HU values, and *p*(*x*| *Θ*) as the *K*-component Gaussian mixture model defined as:

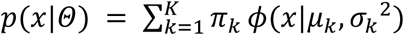

with *Θ* = {*π*_*k*_, *µ*_*k*_, *σ*_*k*_^2^}^*K*^ _*k*=1_, where *π*_*k*_ ≥ *0*, ∑_*k*_ *π*_*k*_ = *1* and *µ*_*k*_, *σ*_*k*_^2^ are the Gaussian mean and variance. We estimated the parameters of the GMM model by minimizing the error in approximating the empirical distribution *p̂*(*x*) and the mixture model *p*(*x*|*Θ*) as:

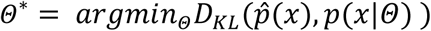

We utilized an expectation-maximization-based approach to estimate the parameters of the GMM model. Once the GMM model parameters *Θ* was estimated, the mean HU of the thymic region (also known as the thymic region for quantification, or TRQ) *A*_*TRQ*_ was estimated as the weighted sum of the individual Gaussian means of the GMM model. Assuming that TRQ consists of two compartments, the adipose and the thymic tissue, each having a characteristic attenuation *A*_*adipose*_ and *A*_*thymic*_, the estimated thymic volume (ETV) was as previously defined^31,38^:

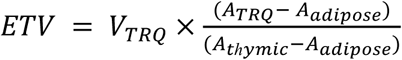

From ETV, we define percent thymic tissue (*pTT*) as:

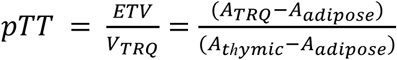

The above formulation ensures *pTT* robustness and invariance under TRQ volume scaling.

### Predictive risk modeling of high-grade pneumonitis

Imaging, clinical, and dosimetric data from the RTOG 0617 and MSKCC cohorts were analyzed. The binary outcome variable considered was the incidence of high-grade (grade ≥ 3) pneumonitis. We evaluated pTT and two dosimetric parameters – volume of the lungs receiving dose ≥ 20 Gy (lung V20) and mean dose to the thymic region (MDTR) – as predictors of risk for high-grade pneumonitis. The analysis was performed for both full cohorts and additionally using median-split subgroups by pTT, lung V20, and MDTR.

### Statistical modeling and risk analysis

We utilized a logistic regression with restricted cubic splines (RCSs) to model potential relationships between each predictor and the binarized outcome of high-grade pneumonitis. RCS offers a flexible tool to improve the model fit, overcoming the limitations of categorical approaches, especially in the case of non-linear associations^42^. Let *x* (pTT, lung V20, or MDTR) be the predictor variables and *y* (the probability of high-grade pneumonitis) be the outcome, then we can model the relationship between *y* and *x* as:

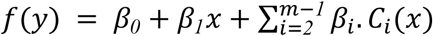

where *f*(. ) is the logit link-function, *C*_*i*_(*x*) is the cubic component in the *i^th^* window, *β_i_* represents the coefficients and *m* represents the number of knots. For *m* knots, the RCS typically has *m* − *1* degrees of freedom for the spline components. The degree of freedom of the model was chosen by minimizing the Bayesian information criterion (BIC) over a set of candidate degrees of freedom values. To generate stable estimates of the coefficients, we utilized the ridge penalized maximum likelihood estimation technique, which was the favored approach given the low event rates. To avoid possible model errors due to outcome imbalance in the dataset, we used the SMOTE oversampling technique within the training folds.

Once the model was fitted, we computed the risk curves, receiver operating characteristics (ROCs), and Fisher’s exact p-value for each of the predictors for the overall cohort as well as the median-split subgroups. The risk curve was obtained by applying the inverse logit function transformation to the fitted RCS model to obtain predicted probabilities across the range of each predictor. ROC-based analysis was used to assess the model’s discriminative characteristics and to derive the area under the ROC curve (AUC) and the Youden optimal cutoff. Fisher’s exact tests were used to assess the association between each predictor and the outcome after dichotomization of the predictors at the Youden optimal cutoff derived from the ROC curves. Five-fold cross-validation was used to obtain robust risk and ROC curves. Additionally, we modeled the probability of high-grade pneumonitis using multivariable logistic regression with restricted cubic splines (RCS) to capture the non linear effects. To obtain the conditional risk curves, we fixed lung V20 values at the lower quartile (Q1), the median (Q2) and the upper quartile (Q3) and computed the probability *Pr*(*Y*|*pTT* = *x*, *V20* = *v*_*j*_) by sweeping *x* across the range of observed pTT values. Here, *y* represents high grade pneumonitis incidence, *V20* being the lung V20 value analyzed at 3 different levels: *v*_*j*_ = {*Q1*, *Q2*, *Q3*}.

## RESULTS

### Clinical cohorts and patient characteristics

We analyzed baseline imaging, dosimetric, and clinical outcomes data for patients in two distinct cohorts: RTOG 0617 (n = 490) and MSKCC (n = 230). Patient characteristics are summarized in **Table 1** and the CONSORT diagram in **Figure 1**. All patients were included in the pTT analyses. After removing cases with missing data, a subset of patients on the RTOG 0617 cohort (n = 459) and (n = 486) was included in the lung V20 and mean thymic dose analyses, respectively. High-grade pneumonitis or radiation pneumonitis (RP) occurred in 28 patients (5%) in RTOG 0617 and in 22 patients (9.56 %) in the MSKCC cohort. Median age was 64 and 68 years for the RTOG 0617 and MSKCC cohorts, respectively. Median overall survival (in months) was 21.9 and 31.2 for the RTOG 0617 and MSKCC cohorts, respectively. **Table 2** summarizes patient characteristics by various subgroups across cohorts.

**Figure 1.**
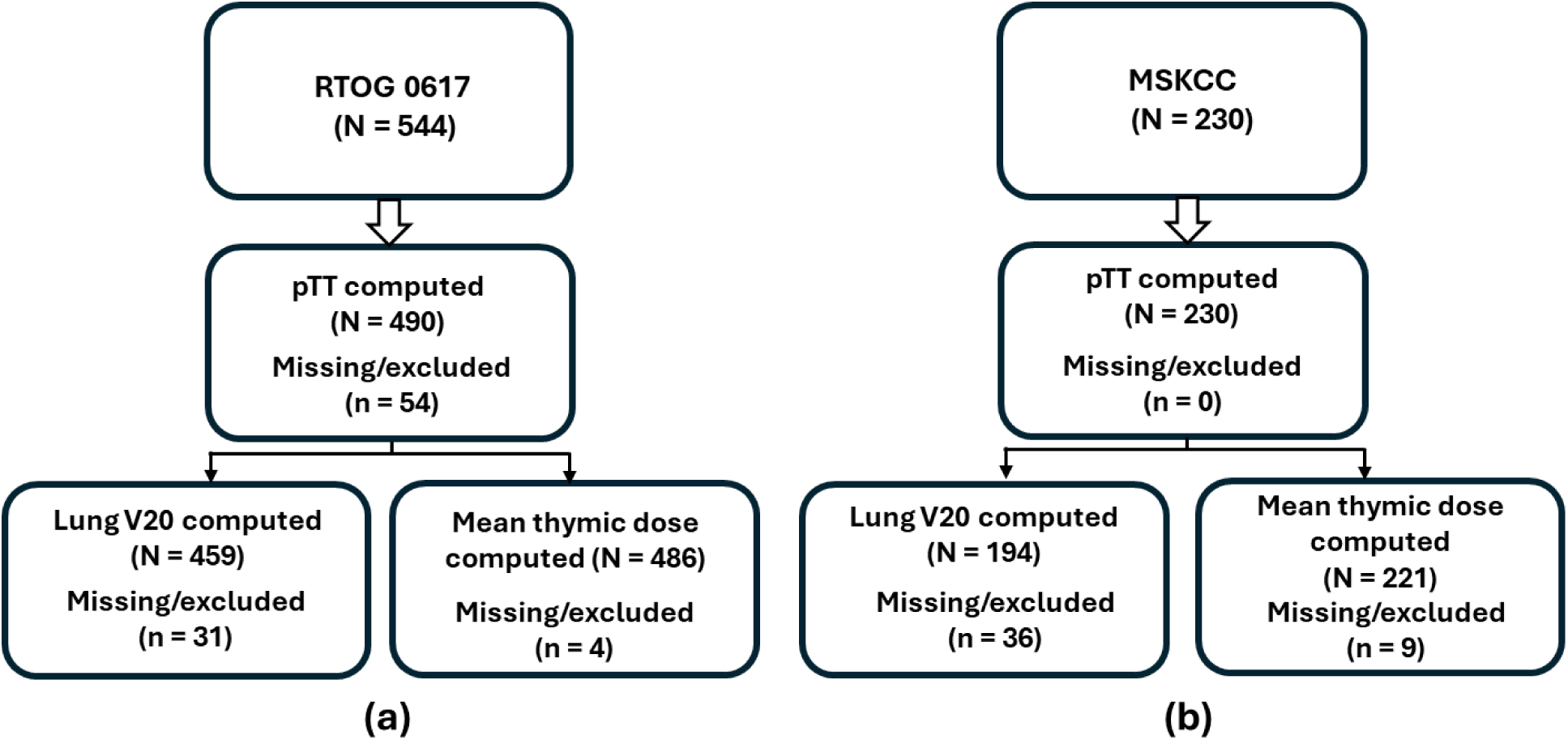
Data distribution across the two cohorts (a) RTOG 0617 cohort-based distribution of the overall number of patients and patients across different subgroups based on pTT, lung V20, and mean thymic dose (and the number of missing/excluded patients within each subgroup). (b) MSKCC cohort-based distribution of the overall number of patients and patients across different subgroups based on pTT, lung V20, and mean thymic dose (and the number of missing/excluded patients within each subgroup).

**Table 1.** Overall patient characteristics across the RTOG 0617 and the MSKCC cohorts.

| Characteristics | MSKCC (N = 230) | RTOG 0617 (N = 544) |
| --- | --- | --- |
| Age: Median (Range) | 68 (45-87) | 64 (37-83) |
| Male: Population (%) | 135 (59%) | 295(60%) |
| Female: Population (%) | 95 (41%) | 200 (40%) |
| AJCC Stage: Population (%) | IIIA: 67 (29%), IIIB: 123 (53%),<br>IIIC: 40 (17%) | IIIA: 324 (60%), IIIB: 171 (31%),<br>NR: 49 (9%) |
| High grade pneumonitis:<br>No. (%) | 22 (10%) | 28 (5%) |

Abbreviations: RTOG, NRG Oncology Radiation Therapy Oncology Group (RTOG) 0617 trial; AJCC, American Joint Committee on Cancer; MSKCC, Memorial Sloan Kettering Cancer Center; NR, not recorded/reported

**Table 2.** Patient characteristics across subgroups within RTOG 0617 and MSKCC cohorts.

| Cohort | Subgroup | Population (N) | Male (%) | Female (%) | High grade pneumonitis incidence |
| --- | --- | --- | --- | --- | --- |
| MSKCC | pTT < Median | 115 | 83 (72%) | 32 (28%) | 16 (14%) |
|  | pTT ≥ Median | 115 | 52 (45%) | 63 (55%) | 6 (5%) |
|  | Lung V20 < Median | 97 | 59 (61%) | 38 (39%) | 4 (4%) |
|  | Lung V20 ≥ Median | 97 | 55 (57%) | 42 (43%) | 15 (15%) |
|  | Mean thymic region dose < Median | 110 | 74 (67%) | 36 (33%) | 11 (10%) |
|  | Mean thymic region dose ≥ Median | 111 | 56 (50%) | 55 (50%) | 11 (10%) |
| RTOG 0617 | pTT < Median | 245 | 159 (69%) | 72 (31%) | 18 (7%) |
|  | pTT ≥ Median | 245 | 112 (49%) | 117 (51%) | 7 (3%) |
|  | Lung V20 < Median | 229 | 138 (60%) | 91 (40%) | 8 (3%) |
|  | Lung V20 ≥ Median | 230 | 133 (58%) | 97 (42%) | 17 (7%) |
|  | Mean thymic region dose < Median | 243 | 141(61%) | 90 (39%) | 10 (4%) |
|  | Mean thymic region dose ≥ Median | 243 | 128 (57%) | 97 (43%) | 15 (6%) |

Abbreviations: pTT, percent thymic tissue; RTOG, NRG Oncology Radiation Therapy Oncology Group; MSKCC, Memorial Sloan Kettering Cancer Center

### Association of pTT with high-grade pneumonitis

Across both studied cohorts, pTT was inversely associated with the incidence and modeled probability of high-grade pneumonitis. In the RTOG 0617 cohort, pneumonitis occurred in 7.3% of patients with pTT less than the median (low pTT group) versus 2.9% with pTT greater than or equal to the median (high pTT group, *p* = 0.038). This association remained consistent after stratification by lung V20: among patients with lung V20 above the median, pneumonitis occurred in 11.5% of those with low pTT versus 2.8% of those with high pTT (p=0.009). Among patients with lung V20 below the median, corresponding rates were 3.7% and 3.3%, respectively (p=0.49). Thus, patients with both high lung V20 and low pTT represented the subgroup at greatest risk (11.5%, n = 14), whereas those in all other other subgroups had few events (no more than 4 recorded events and at most 3.7%) (**Figure 2**).

**Figure 2.**
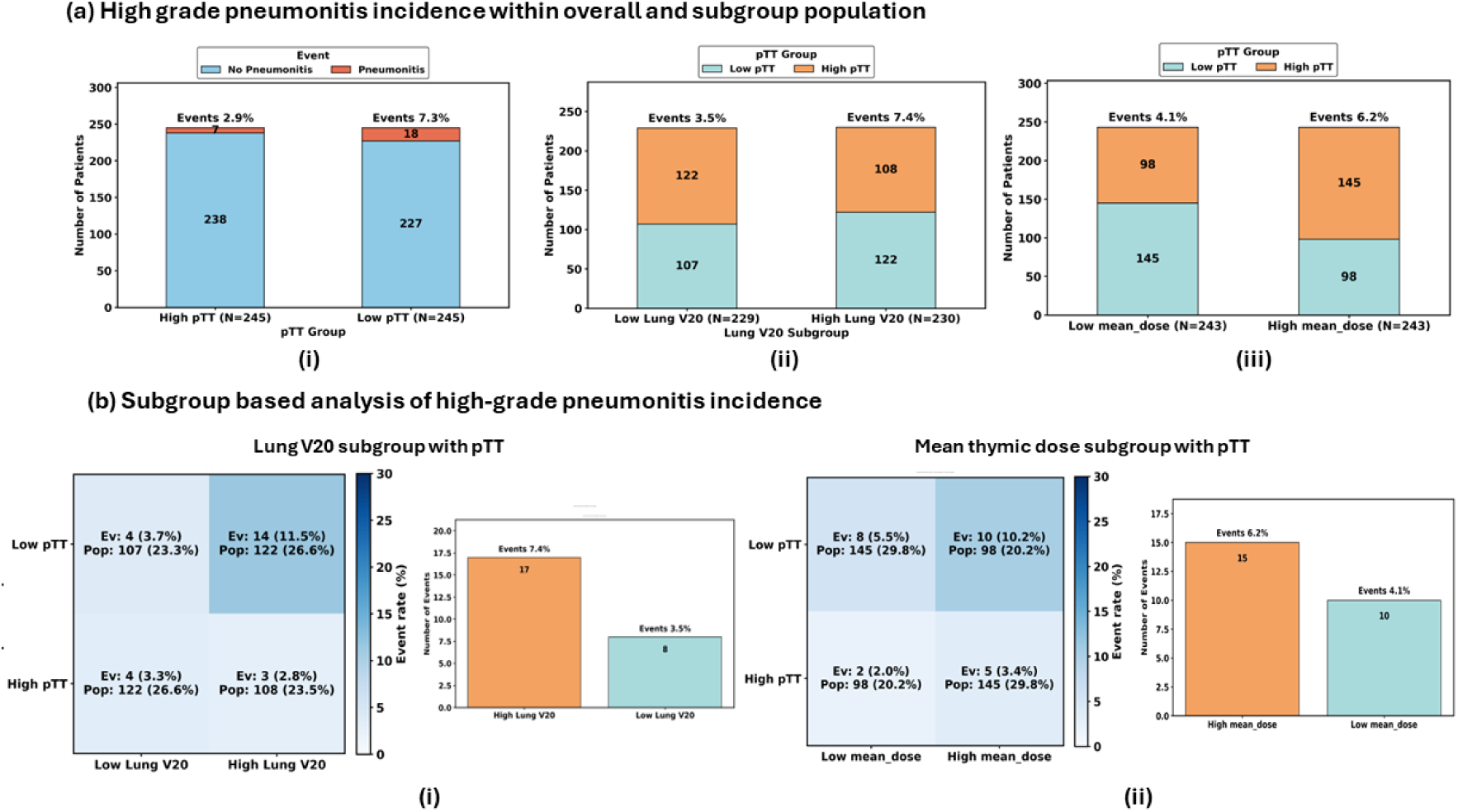
High-grade pneumonitis incidence distribution within the overall RTOG 0617 cohort and the median-split Lung V20 and Mean Thymic Dose subgroups. (a) High-grade pneumonitis incidence in the overall cohort as well as subgroups: (i) pTT subgroup (ii) Lung V20 subgroup (iii) Mean thymic dose subgroup (b) Subgroup-based pneumonitis incidence analysis with pTT: (i) pTT vs Lung V20, (ii) pTT vs Mean thymic dose. The low pTT and high pTT groups had pneumonitis incidence of 7.3% vs. 2.9%, p = 0.038. The low lung V20 and high lung V20 groups had pneumonitis incidences of 3.5% vs 7.4%, p = 0.16. The high mean thymic dose and low mean thymic dose subgroups had pneumonitis incidence of 6.2% vs 4.1%, p = 0.41. The p-values are calculated using the median cutpoint.

Among patients treated at MSKCC, the incidence of high-grade pneumonitis was 13.9% (n = 16) versus 5.2% (n = 6) (*p* = 0.042) for the low and high pTT subgroups, respectively. This pattern persisted after stratification by lung V20: among patients with a lung V20 below the median, high-grade pneumonitis incidence was 6.8% in those with low pTT compared to 1.9% of those with high pTT (p=0.016), whereas among those with a lung V20 above the median, the corresponding rates were 20.8% and 9.1% respectively (0.022). Therefore, the combination of high lung V20 and low pTT represented the subgroup with the highest rate of severe pneumonitis (20.8%, n = 11), whereas events were infrequent among patients with low lung V20 and high pTT (1.9%, n = 1)^31^.

Modeled risk curves demonstrated a monotonic decrease in pneumonitis probability with increasing pTT, with narrow 1-SD confidence bands confirming the downward trend across RTOG 0617 (**Figure 4**) and MSKCC (**Figure 5**) cohorts. Five-fold cross-validation yielded mean ROC AUCs of 0.60 for the RTOG cohort and 0.64 for the MSKCC cohort, indicating modest yet consistent discriminative ability above chance. When the model’s predicted risk was dichotomized using the Youden optimal cutoff, a statistically robust association between model-derived risk and pneumonitis events was observed (*p* = 0.003), reinforcing pTT’s value as an independent predictor of high-grade pneumonitis. Figures 4 and 5 depict the modeled risk curves and ROC analyses for pTT, lung V20, and MDTR.

### Association of lung V20 with high-grade pneumonitis

Across both cohorts, higher lung V20 was consistently associated with an increased incidence of high-grade pneumonitis. In the RTOG 0617 cohort, pneumonitis occurred in 7.4% of patients with high lung V20 compared with 3.5% in those with low lung V20 (*p* = 0.16) (**Figure 2**). In the MSKCC cohort, the corresponding rates were 15.5% and 4.1%, respectively (*p* = 0.014) (**Figure 3**).

**Figure 3.**
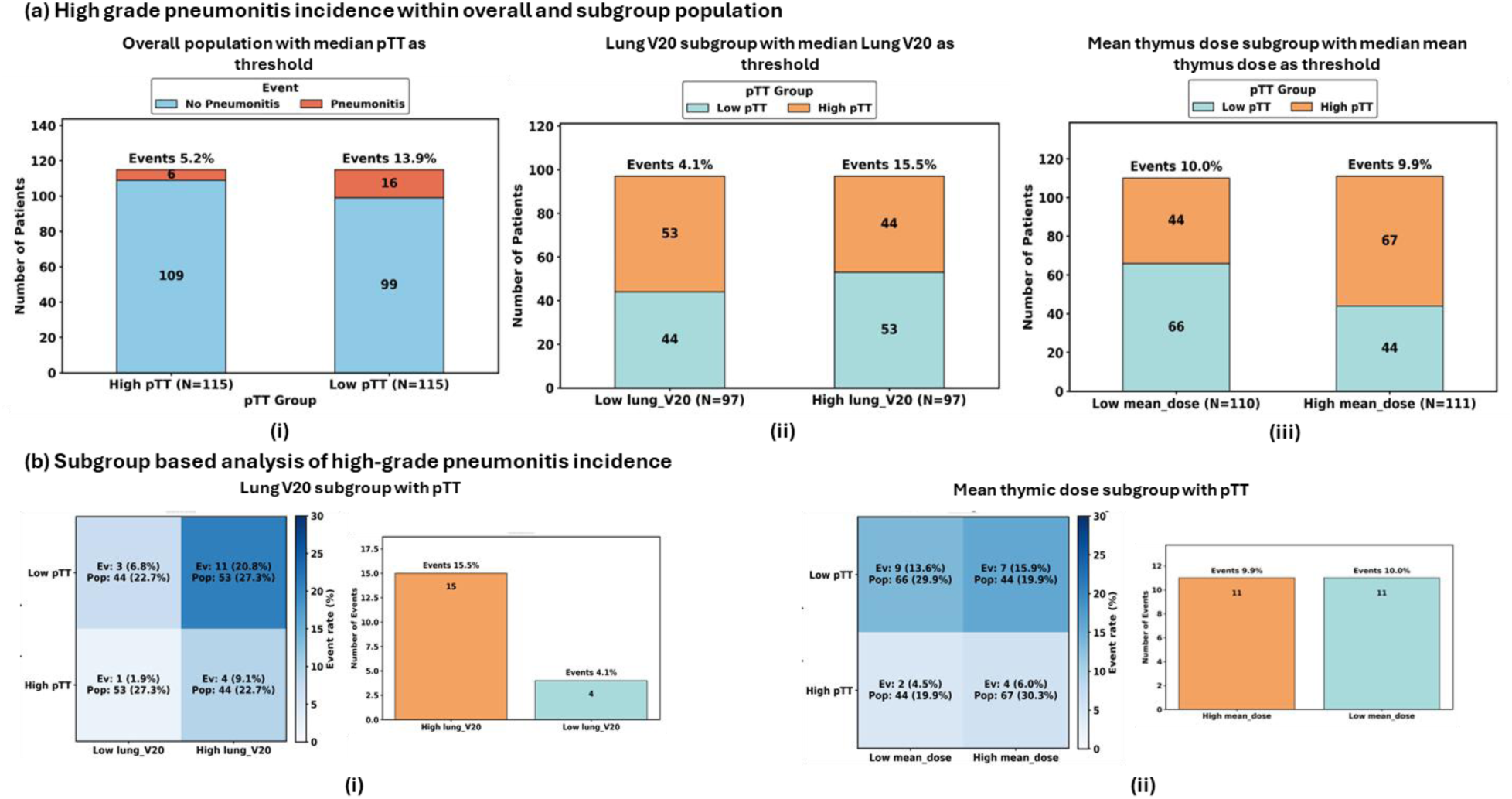
High-grade pneumonitis incidence distribution within the overall MSKCC cohort and the median-split Lung V20 and mean thymic dose subgroups. (a) High-grade pneumonitis incidence in the overall cohort as well as subgroups: (i) pTT subgroup (ii) Lung V20 subgroup (iii) Mean thymic dose subgroup (b) Subgroup-based pneumonitis incidence analysis with pTT: (i) pTT vs Lung V20, (ii) pTT vs Mean thymic dose. The low pTT and high pTT groups had pneumonitis incidences of 13.9% vs 5.2%, p = 0.042. The low lung V20 and high lung V20 groups had pneumonitis incidences of 4.1% vs 15.5%, p = 0.014. The high mean thymic dose and low mean thymic dose subgroups had pneumonitis incidence of 9.9% vs 10.0%, p = 1. The p-values are calculated using the median cutpoint.

Univariate analysis demonstrated a monotonic increase in pneumonitis probability with rising lung V20 across both cohorts (**Figure 4 and 5**). Mean cross-validated AUCs were 0.61 for RTOG 0617 and 0.79 for MSKCC, indicating good discriminatory performance for identifying patients at risk. Using the Youden optimal cutoff, Fisher’s exact test yielded *p* = 0.023 and *p* = 0.0008 for RTOG 0617 and MSKCC, respectively, confirming, as expected, a statistically robust association between lung V20 and high-grade pneumonitis risk.

**Figure 4.**
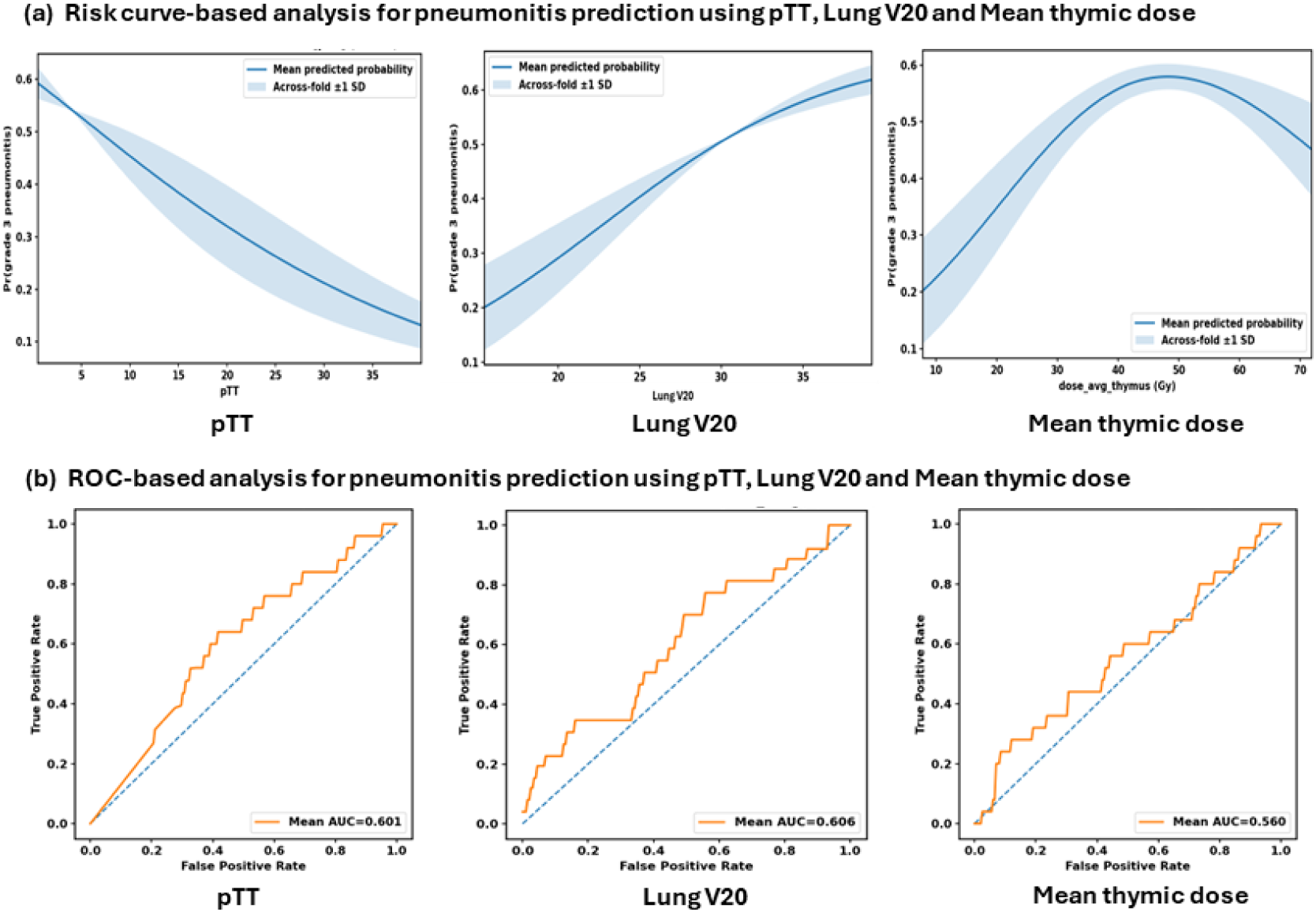
Risk curves and ROC-based analysis of high-grade pneumonitis prediction for the RTOG cohort using pTT (Fisher p-value: 0.022), Lung V20 (Fisher p-value: 0.023), and mean thymic dose (Fisher p-value: 0.101) as predictors. (a) Risk curves computed for the predictors. (b) ROC-based analysis. Fisher p-values are calculated based on the model using the Youden cutoff from the ROC-based analysis.

**Figure 5.**
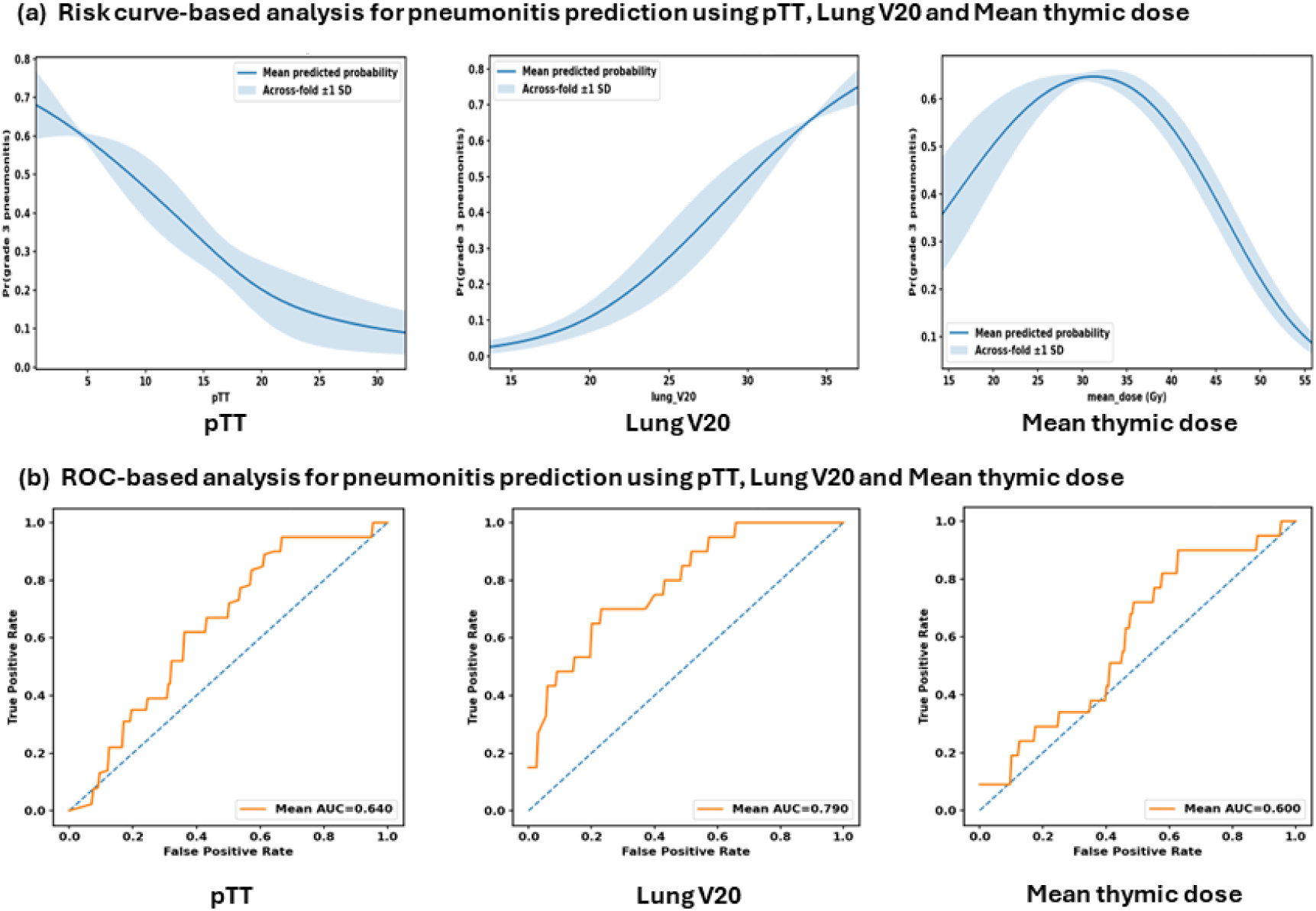
Risk curves and ROC-based analysis of high-grade pneumonitis prediction for the MSKCC cohort using pTT (Fisher p-value: 0.003), Lung V20 (Fisher p-value: 0.0008), and mean thymic dose (Fisher p-value: 0.015) as the predictors. (a) Risk curves computed for the predictors. (b) ROC-based analysis. Fisher p-values are calculated based on the model using the Youden cutoff from the ROC-based analysis.

### Association of mean thymic dose with high-grade pneumonitis

In the RTOG 0617 cohort, pneumonitis occurred in 6.2% of patients receiving high MDTR compared with 4.1% in the low-dose group (*p* = 0.411), a nonsignificant numerical difference (**Figure 2**). Similarly, in the MSKCC cohort, pneumonitis incidence did not differ by MDTR (9.9% for high MDTR vs. 10.0% for low MDTR groups, *p* = 1.0) (**Figure 3**).

Modeled analysis revealed a nonlinear relationship between MDTR and predicted pneumonitis risk; risk increased with dose up to a threshold, then declined at higher values (**Figure 4 &5)**. Cross-validated ROC analysis demonstrated modest discrimination, with mean AUCs of 0.56 for RTOG 0617 and 0.60 for MSKCC. Fisher’s exact test using the Youden optimal cutoff yielded *p* = 0.10 and *p* = 0.015, respectively. These results suggest that MDTR had a weaker and less consistent association with high-grade pneumonitis compared to pTT or lung V20.

Stratification by baseline pTT revealed that MDTR predicted pneumonitis risk among patients with low pTT (10.2% for high MDTR vs. 5.5% for low MDTR groups, p = 0.21). The combination of high MDTR and low pTT was associated with the highest pneumonitis incidence (10.2%, *n* = 10), while high pTT with low MDTR identified the lowest-risk subgroup (2.0%, *n* = 2); Fisher p value = 0.0326.

### Conditional risk analysis

Conditional risk curves were generated by fixing lung V20 at the lower quartile (25% for RTOG 0617 and 22% for MSKCC), median (30% and 28%), and upper quartile (35% and 34%) within each cohort, while varying pTT across its observed range to estimate the probability of high-grade pneumonitis. As demonstrated in **Figure 6**, pneumonitis risk decreased progressively with increasing pTT across all lung V20 strata for both the RTOG 0617 and MSKCC cohorts. Overall risk was greatest at higher lung V20 values and lowest at lower lung V20 values. Notably, even among patients with high lung V20, higher pTT values were associated with a substantially lower risk of pneumonitis compared with lower pTT values, indicating a consistent protective effect of thymic composition independent of lung dose.

**Figure 6.**
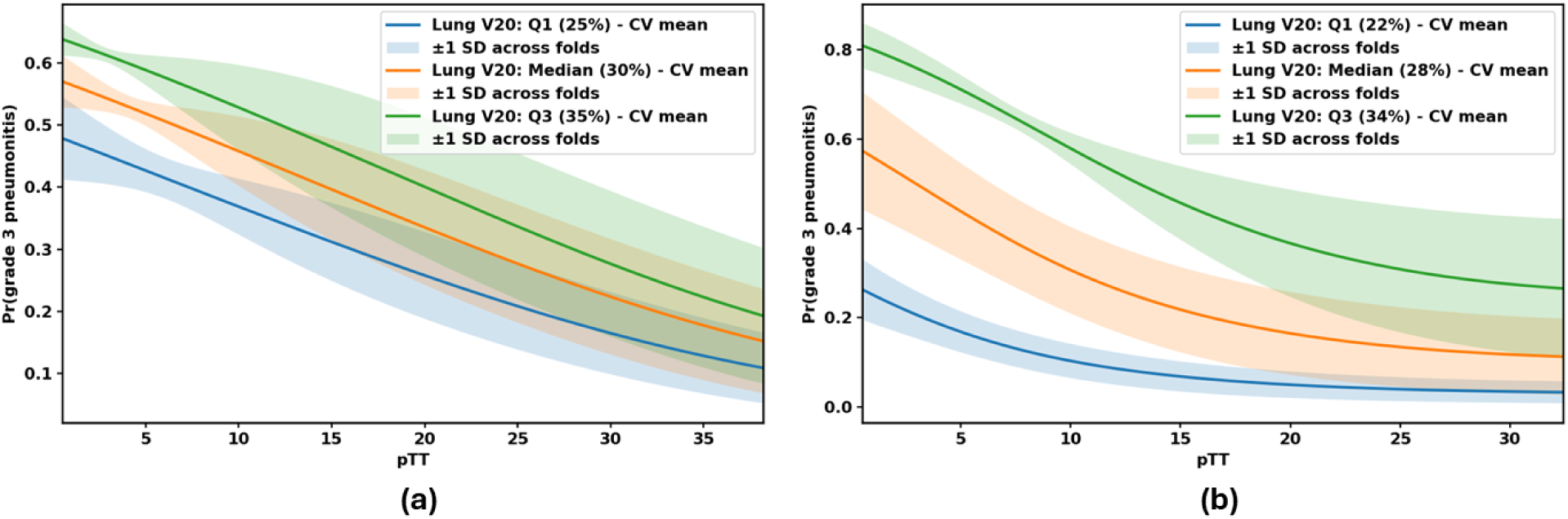
Multi variable risk analysis of high grade pneumonitis incidence as a function of pTT (continuous predictor) and Lung V20 (fixed at 3 different levels of Lung V20: Lower quartile (Q1), Median (Q2) and upper quartile (Q3)) across the (a) RTOG 0617 cohort (b) MSKCC cohort. The curve shows decreased high grade pneumonitis risk with increase in pTT and is stratified by the level of Lung V20. The solid lines indicates the mean cross validated (CV) risk, while the shaded area denotes ±*1* standard deviation (SD) across different folds.

## DISCUSSION

This study establishes percent thymic tissue (pTT)—a radiographic measure of residual functional thymic tissue in adults—as a novel, independent predictor of severe pneumonitis in patients with locally advanced NSCLC treated with cCRT, with or without consolidation immunotherapy. Patients with low baseline pTT experienced a significantly higher incidence of high-grade pneumonitis, with risk decreasing progressively as pTT increased. This effect persisted after stratification by lung radiation dose, indicating that the association between low pTT and pneumonitis risk was independent of established lung dosimetric factors ^10,11^. The association was also consistent across treatment eras, observed in both the RTOG 0617 cohort treated with cCRT alone and the MSKCC cohort treated according to the modern standard of care, which incorporates consolidation durvalumab per PACIFIC ^1–3,6,7^. Importantly, patients with a low baseline pTT who were also exposed to high lung radiation doses in either cohort demonstrated the highest frequency of severe pneumonitis events, identifying a particularly vulnerable subgroup requiring intensive monitoring and potentially modified treatment approaches. While radiation dose to the thymic region alone was not independently associated with pneumonitis risk, among patients with low baseline pTT, higher mean thymic dose significantly increased risk for severe pneumonitis. This finding suggests that patients with limited thymic reserve may be more susceptible to treatment-related immune injury, reflecting impaired capacity for homeostatic immune reconstitution post-cCRT. Overall, low pTT was associated with an increased risk of pneumonitis, whereas high pTT appeared to be protective.

The mechanisms underlying these observations are likely multifactorial. The protective association of high pTT with reduced risk of severe pneumonitis likely reflects the role of preserved thymic function in maintaining immune homeostasis during recovery from chemoradiation. cCRT induces profound immunosuppression, and the convalescent phase requires coordinated regeneration of effector and regulatory T-cell compartments^7,43^. Patients with higher pTT likely retain greater thymic capacity to restore regulatory T-cell (Treg) populations, thereby constraining excessive inflammatory rebound during immune reconstitution. In the context of checkpoint blockade, this balanced recovery may temper effector-cell (CD8⁺, Th1) hyperactivation within the lung parenchyma, reducing the likelihood of uncontrolled immune-mediated lung injury. This interpretation aligns with prior literature demonstrating that preserved thymic integrity supports both effective antitumor immunity and protection against autoimmune or inflammatory toxicity when immune checkpoints are inhibited^44–46^. Irradiation of the thymic region may blunt this protective effect by depleting Treg progenitors and, at higher doses, damaging the epithelial and mesenchymal scaffolds required for T-cell maturation. The increased incidence of pneumonitis observed in patients receiving high thymic doses, particularly among those with low pTT, supports this mechanism and suggests that thymic irradiation may exacerbate vulnerability in patients with limited baseline reserve. Nevertheless, although thymic radiation dose may contribute to immune perturbation, baseline thymic function, reflected by pTT, appears to be the dominant determinant of immune resilience and the primary driver of the observed association with severe pneumonitis.

Mean thymic radiation dose demonstrated a more complex, non-linear association with pneumonitis risk compared to pTT and lung V20. The risk curves showed an initial increase in pneumonitis probability with rising thymic dose, followed by a decline at the highest dose levels. This pattern likely reflects competing biological processes. At moderate thymic doses, partial injury to thymic epithelium and progenitor niches may impair regulatory T-cell (Treg) renewal while leaving sufficient effector capacity intact, thereby predisposing to dysregulated immune reconstitution and heightened inflammatory susceptibility. However, at very high doses, near-complete ablation of thymic activity may blunt both effector and regulatory regeneration, attenuating immune reconstitution altogether and paradoxically reducing the likelihood of clinically apparent immune-mediated pneumonitis. Such dose-dependent biphasic relationships have been described in other immune contexts, where intermediate damage promotes dysregulated inflammation while more profound suppression dampens immune activation^47–49^. In this framework, baseline thymic reserve (pTT) modulates the slope of this relationship; patients with low pTT entering treatment with limited reserve may cross the threshold for immune dysregulation at substantially lower radiation doses, thereby predisposing them to severe pneumonitis.

Pneumonitis is a dose-limiting toxicity that, in the modern treatment era, not only causes substantial morbidity but also frequently renders patients ineligible for, or leads to interruption or discontinuation of, potentially life-prolonging immunotherapy^1,2,7^. The PACIFIC, PACIFIC-2, and CheckMate 73L trials established the role and sequencing of immunotherapy in the management of unresectable stage III NSCLC, but they also underscored new challenges in balancing efficacy and toxicity, particularly with respect to eligibility for consolidation immunotherapy ^1–3,6,7^. In PACIFIC, patients were eligible for durvalumab only if they had completed cCRT without early progression, had recovered from cCRT-related toxicities to ≤ grade 2, and maintained good performance status ^1–3^. By contrast, PACIFIC-2 enrolled patients prior to cCRT initiation, before it was known which patients would tolerate or respond to therapy. As a result, PACIFIC-2 inherently included individuals who, in PACIFIC, would have been among the 27–50% of patients ineligible for consolidation durvalumab because of early progression or treatment-related toxicity ^1,7^. Real-world treatment patterns more closely mirror the all-comer population represented in PACIFIC-2, which had less favorable outcomes, notwithstanding the limitations of cross-trial comparisons. This observation underscores a critical need to better understand and predict toxicity within contemporary multimodality regimens, as real-world populations may not be realizing the full benefit observed in PACIFIC. Our results suggest that baseline thymic function, quantifiable using percent thymic tissue (pTT) on standard pretreatment CT scans, may help identify patients at elevated risk for these complications, informing both patient selection and individualized toxicity-mitigation strategies. The identification of pTT as a predictive biomarker for severe pneumonitis enables improved risk stratification using standard pre-treatment CT scans. Patients with low pTT could be identified prospectively as high-risk candidates who may benefit from optimized and more stringent lung dose constraints and closer clinical monitoring during and after treatment. In the current study, we provide corresponding risk estimates stratified by lung V20, enabling clinicians to map individual patients treated per the current standard of care onto specific risk trajectories based on their baseline pTT and modifiable dosimetric profile (Figure 6B).

This study has several limitations, including its retrospective design and the relatively low number of pneumonitis events in certain subgroups, which limited statistical power. In particular, the small number of events constrained our ability to formally evaluate the predictive power of pTT while controlling for lung V20. Nevertheless, in both cohorts, patients with high lung V20 had significantly higher pneumonitis rates when pTT was low versus high. Among patients with low lung V20, this relationship remained statistically significant in the MSKCC cohort and trended similarly in the RTOG cohort (p = 0.49, likely reflecting the limited number of events). The consistency of these findings across cohorts and lung V20 strata supports the robustness of pTT as an independent predictor of pneumonitis, warranting validation in larger datasets. The scope of analysis was also restricted to pneumonitis, excluding other treatment-related toxicities such as cardiac, esophageal, neurologic, and hematologic injury. However, this focus is justified by both clinical relevance and radiobiological plausibility. Based on first principles of radiation oncology, the lungs behave as a parallel organ, in which toxicity reflects the cumulative effect of injury across a distributed parenchymal volume^50–52^. In contrast, structures such as the esophagus, spinal cord, and coronary vessels are serial organs, where toxicity is driven primarily by focal dose metrics and immune modulation plays a modest role. Any apparent associations in such organs would likely represent an artifact of direct tissue injury from localized high-dose exposure rather than systemic immune effects. On the other hand, because radiation-induced lung damage is distributed and sublethal at the cellular level, immune and inflammatory mechanisms play a more prominent role in determining whether local injury resolves or propagates^53,54^. Accordingly, immune-mediated variation in radiation sensitivity is more biologically relevant for lung injury, making the lungs a particularly appropriate focus for investigating host-specific determinants such as thymic reserve.

In conclusion, this study establishes a significant link between pTT, a novel measure of thymic composition, and risk for severe pneumonitis in locally advanced NSCLC. Our results suggest a robust thymic reserve is protective against severe pulmonary toxicity, especially in the modern immunotherapy era. While lung dose remains a powerful predictor of radiation-induced lung injury and pneumonitis, pTT provides complementary, host-specific biological information to enhance prediction and mitigation of high-grade pneumonitis and optimization for consolidation in the immunotherapy era. Ongoing studies are validating pTT using comprehensive immune profiling and large-scale genomic datasets uniquely available at MSKCC to elucidate the molecular and immunologic basis of the observed associations. Future smart clinical trial designs may incorporate pTT, upon further validation, into multimodal risk stratification frameworks to optimize both disease control and toxicity management.

## Data Availability

The study was conducted using an MSKCC institutional cohort of radiotherapy patients obtained with IRB approval, and a publicly available dataset, available on TCIA: https://www.cancerimagingarchive.net/collection/nsclc-cetuximab/

## Data availability

This study utilized two datasets comprising a total 720 patients who met inclusion criteria for analysis. The first was obtained from the NCTN/NCORP Data Archive of the National Cancer Institute (NCI). These data were originally collected from clinical trial NCT00533949 (RTOG-0617) and are publicly available for download with a Data Use Agreement (DUA) through The Cancer Imaging Archive (TCIA): https://www.cancerimagingarchive.net/collection/nsclc-cetuximab/

In addition, an institutional cohort of radiotherapy patients treated at Memorial Sloan Kettering Cancer Center (MSKCC) was analyzed under an Institutional Review Board (IRB)-approved protocol. These data contain protected health information and are not publicly available due to institutional and patient privacy restrictions.

## ACKNOWLEDGEMENTS

This manuscript was prepared using data from Datasets NCT00533949 from the NCTN/NCORP Data Archive of the National Cancer Institute’s (NCI’s) National Clinical Trials Network (NCTN), NCTN/NCORP. Data were originally collected from clinical trial NCT number NCT00533949 (Study identification: RTOG-0617). All analyses and conclusions in this article are the sole responsibility of the authors and do not necessarily reflect the opinions or views of the clinical trial investigators, the NCTN, the NCORP or the NCI. The authors gratefully acknowledge NRG Oncology/RTOG and the principal investigator of RTOG 0617, Dr. Jeffrey Bradley, for making the trial data publicly available. Their commitment to data sharing has enabled this secondary analysis and supports continued progress for patients affected by lung cancer. The Principal Investigator of the current analysis (TC) assumed responsibility to lead the scientific investigation presented using the data.

## Code Availability

Source code for the pTT computation pipeline is available upon reasonable request to the corresponding author and will be openly released following peer review.

## SUPPLEMENTAL RESULTS

### Subgroup-Based Analysis of High-Grade Pneumonitis Incidence

Subgroup analyses were performed to evaluate the association between pTT and high-grade pneumonitis across strata defined by lung V20 and mean thymic dose (median split).

In both cohorts, pTT retained its inverse association with pneumonitis risk across all subgroups. In the MSKCC cohort, risk curves demonstrated a pronounced decline in pneumonitis probability with increasing pTT, particularly within the low lung V20 subgroup. Patients with low lung V20 exhibited sharply reduced risk at higher pTT values, whereas risk remained elevated in the high lung V20 subgroup. ROC analysis showed stronger discriminatory ability for pTT in the low lung V20 subgroup (mean AUC = 0.80) than in the high lung V20 subgroup (mean AUC = 0.64). Fisher’s exact test using the Youden optimal cutoff yielded p = 0.0215 for the high-V20 subgroup and p = 0.016 for the low-V20 subgroup.

Similar trends were observed in the RTOG cohort, though limited event counts in the low-V20 subgroup reduced statistical robustness. For RTOG, Fisher’s test produced p = 0.00915 for the high-V20 subgroup and p = 0.4908 for the low-V20 subgroup, with corresponding AUCs of 0.632 and 0.5160.

When stratified by mean thymic dose, ROC analyses yielded mean AUCs of 0.65 and 0.56 for the high- and low-dose subgroups, respectively. Fisher’s exact test produced p = 0.007 and p = 0.192, confirming a stronger and more consistent association between pTT and pneumonitis risk in the high-dose subgroup.

## SUPPLEMENTAL FIGURES

**Figure S1.**
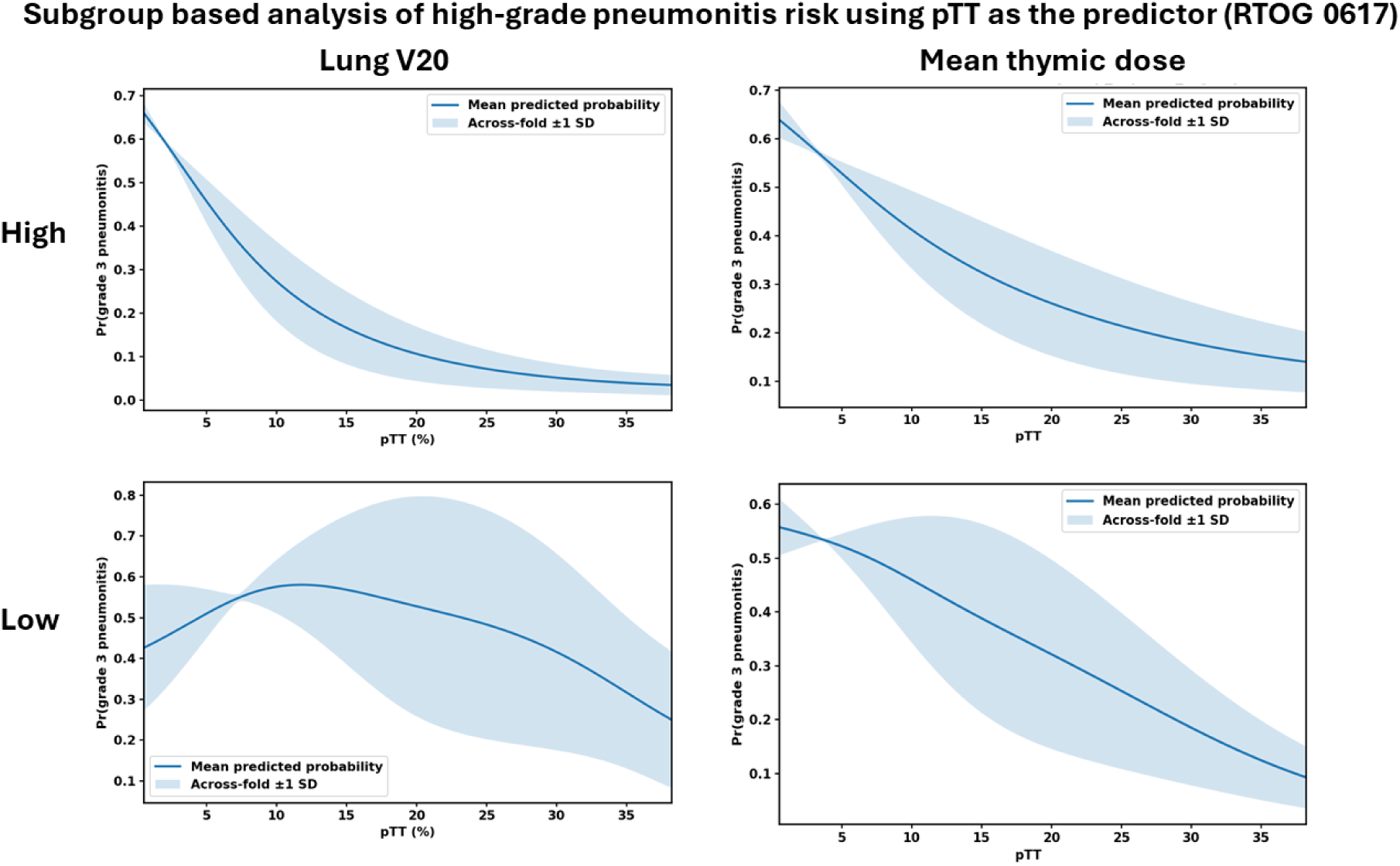
Risk curves analysis of high-grade pneumonitis for high-low lung V20 and mean thymic dose subgroups within the RTOG cohort using pTT. For the high lung V20 subgroup, Fisher’s exact test yielded a p-value of 0.009, while for the low lung V20 subgroup, the p-value obtained was 0.49. For the mean thymic dose, the high and the low subgroups yielded a p-value of 0.0074 and 0.192, respectively. Fisher p-values are calculated based on the model using the Youden cutoff from the ROC-based analysis

**Figure S2.**
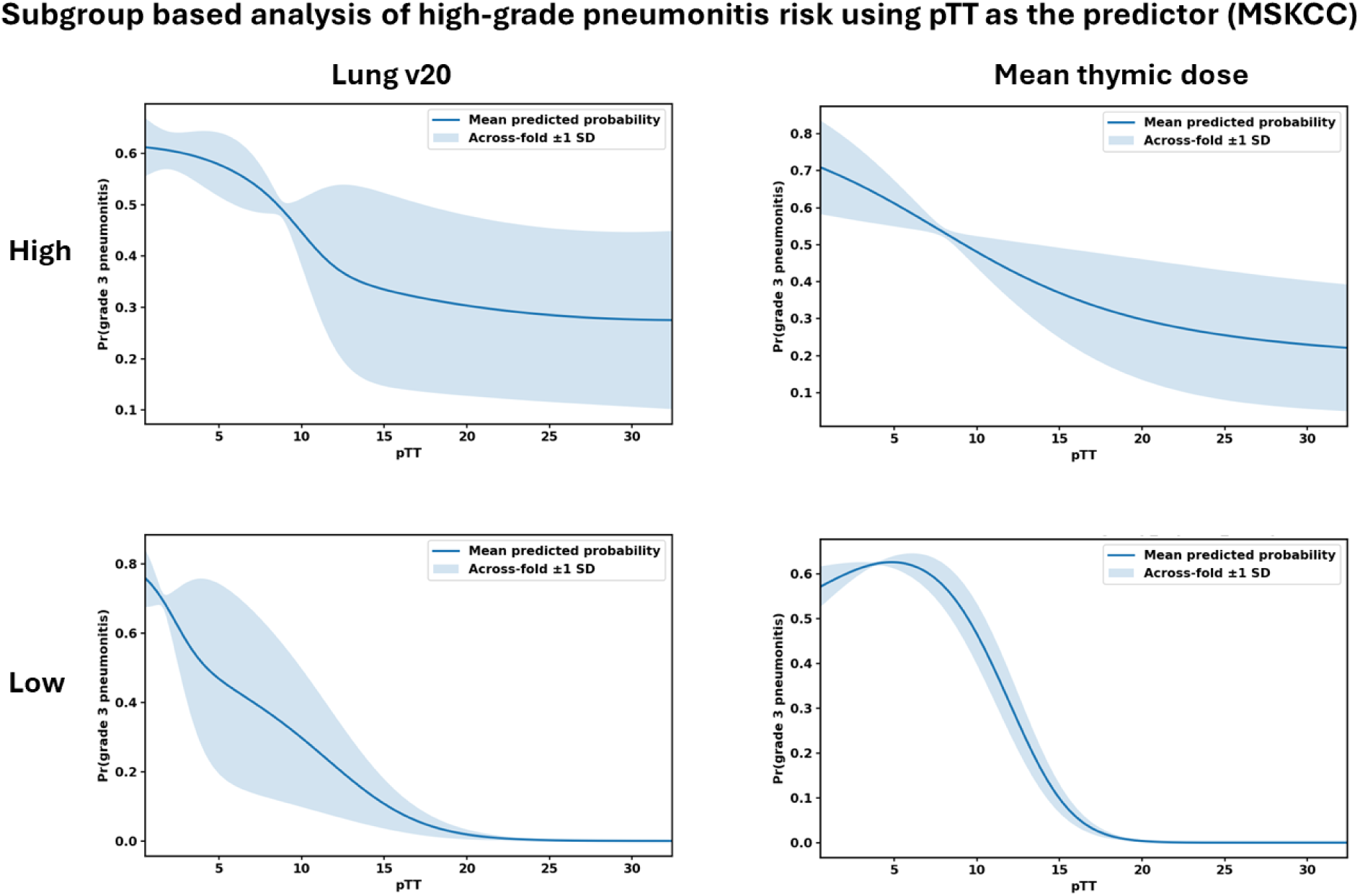
Risk curves analysis of high-grade pneumonitis for high-low lung V20 and mean thymic dose subgroups within the MSKCC cohort using pTT. For the high lung V20 subgroup, Fisher’s exact test yielded a p-value of 0.022, while for the low lung V20 subgroup, the p-value obtained was 0.016. For the mean thymic dose, the high and the low subgroups yielded a p-value of 0.033 and 0.015, respectively. Fisher p-values are calculated based on the model using the Youden cutoff from the ROC-based analysis.

